# A Systematic Review and Global Meta-analysis of Secondary Fungal Infections Associated with COVID-19

**DOI:** 10.1101/2024.10.25.24316125

**Authors:** Aravind Murugavel, Sridevi Raghunathan, Jayapradha Ramakrishnan

## Abstract

**Background:** The COVID-19 pandemic has exposed patients to severe secondary fungal infections, exacerbating clinical outcomes and devastating impact. This study conducts a systematic review with meta-analysis of secondary fungal infections (SFIs) associated with COVID-19 considering various significant parameters, such as the frequency of SFIs across the globe, species shift, gender-specific infection rates, the significance of medical history, efficacy of steroid and antifungal, treatment outcomes (mortality rate), and fungal- polymicrobial mortality analysis.

**Methods:** A literature search was conducted on COVID-19-related fungal infection studies (2020–2024) from SCOPUS and PUBMED databases, excluding preprints. The systematic data extraction captured the PMCID, country, patient demographics (age and gender), clinical outcomes, associated pathogens, medical history, and treatment details.

**Findings:** The global meta-analysis of COVID-19-associated SFIs yielded 10,700 cases across 58 countries, exhibiting a significant male predominance (65.6% vs. 34.3% female). *Aspergillus* spp., *Candida* spp., and *Mucorales* spp. emerged as the primary fungal pathogens. The predominant six countries marking 80 % of global cases include India (46.8 %), Italy (10 %), Iran (9.4 %), France (5.1 %), Spain (4.3 %), and Egypt (4.1 %). Complication rates revealed CAM as the most prevalent (59.2%), with a 28% mortality rate. CAC (21.6%) and CAPA (19.1%) had substantially higher mortality rates, at 54% and 58%, respectively. Specific populations were highly affected, including individuals with diabetes were prone to CAM, those undergone catheterization were at increased risk of CAC, and individuals with respiratory diseases or without prior medical history were susceptible to CAPA. In both CAC and CAPA, the species shift towards the non-albicans spp. and non-fumigatus spp., associated with higher mortality. In addition, polymicrobial infection with fungal pathogens (*Aspergillus* spp., *Candida* spp., *Mucorales* spp.) and Multi-bacteria ( *K. pneumoniae, P. aeruginosa, E. coli, S. aureus*) also increased the mortality rate. Effective treatments were identified, including combining caspofungin with corticosteroids for CAC, voriconazole with dexamethasone for CAPA, and AmBisome monotherapy for CAM.

**Interpretation:** In SFIs populations, CAM prevailed in high-density areas with relatively lower mortality rates, whereas CAC and CAPA exhibited higher mortality rates. Notably, polymicrobial infections significantly increased mortality across all SFIs. Underlying medical conditions primarily influenced the type of fungal pathogen, but treatment outcomes varied. Azole drugs and Amphotericin-B were ineffective against Candidiasis, except for caspofungin’s limited susceptibility. Voriconazole and AmBisome demonstrated efficacy against Aspergillosis and Mucormycosis, respectively. Additionally, steroid administration proved life-saving in CAPA and CAC cases, yet remained ineffective in CAM.

**Funding:** None

**Research in context:** *Evidence before this study:* Five years after the COVID-19 pandemic, a plethora of research has investigated SFIs associated with COVID-19, with a major focus on pathogenicity, immunomodulation, and the impact of steroids and tocilizumab. However, a critical knowledge gap persists that addresses meta-analysis on the frequency of SFIs by country, gender-specific infection rates, the significance of medical history, species shift within the fungal kingdom, its virulence expression, polymicrobial infection dynamics, synergistic effects of steroids and antifungals in this context remains understudied. To address these gaps, the meta-analysis comprehensively examines these critical aspects, shedding light on various aspects of SFIs.

*Added value of this study:* The systematic meta-analysis of COVID-19-associated SFIs revealed the emergence of non-candida spp. and non-fumigatus spp. Polymicrobial infection has been linked to alarming outcomes resulting in a 100% mortality rate.Similarly, the co-infection of *C. albicans* with non-albicans spp. *A. fumigatus* with non-fumigatus spp. increased the mortality rate to 100%. For other species-related, effective therapies such as the combination of caspofungin and corticosteroids against Candidiasis (CAC), voriconazole and dexamethasone (CAPA), and AmBisome monotherapy (CAM) to combat SFIs.

*Implications of all the available evidence:* The geographic distribution of fungal pathogens varies globally, with differing mortality rates. Deciphering their genomic characteristics will unveil insights into behaviour, transmission, and virulence, enabling targeted diagnostics, treatments, and prevention strategies.

**Methodology:** *Data collection:* The peer-reviewed published case studies, multicentric studies, retrospective studies, single-center studies and cohort studies represented with individuals case files were collected using search keywords “COVID-19 and Aspergillus”, COVID-19 and Candida“, COVID-19 and Mucorales”, “COVID Associated Pulmonary Aspergillosis”, “COVID Associated Mucormycosis”, COVID Associated Candidiasis”, in SCOPUS and PUBMED databases. Based on the search results, the articles from Aug 2020 to May 2024 were filtered excluding the preprint articles. A total of 1981 articles that included duplicates, articles unrelated to the study as well without abstracts were eliminated. A systematic review yielded 663 eligible publications, which were subjected to independent individual case meta-analysis. The distribution included 154 studies on CAC, 240 on CAPA, and 269 on CAM **(Figure 1)**. From each article, the details of PMCID, country, age, gender, treatment outcome (live/dead), pathogens, medical history, and usage of steroids, antibacterial & antifungal were systematically collected. For the global surveillance of COVID-19-associated SFIs, the information from review, cohort, and retrospective studies was included.

*Meta-analysis and its statistics:* A systemic global survey was conducted for the 58 reported countries with COVID- 19-associated SFIs in accordance with PRISMA guidelines. The meta-analysis surveillance was conducted considering various significant parameters, such as frequency of SFIs across the globe, species shift, gender-specific infection rates, the significance of medical history, efficacy of steroid and antifungal, treatment outcomes (mortality rate), and fungal- polymicrobial mortality analysis. For the statistical analysis, Jamovi v2.6.2 tool and other tools were used as given below. The proportional meta-analysis was studied using a random effects model to quantify the distribution of SFIs attributed to *Mucorales* spp., *Candida* spp., and *Aspergillus* spp., across the countries. The analysis was considered with a 95% confidence interval (Cl). The analysis also evaluated the overall effect size and sample heterogeneity as indicated by the I² statistic. To investigate the frequency distribution of individual species within SFI, a ClinicoPath table one was employed. This statistical approach enabled the examination of the frequency range of specific species in respective SFIs. To estimate the mortality rates, a two-outcome proportion test was conducted, providing proportion values accompanied by 95 % CI. This study calculated the overall respective SFIs ( CAM , CAPA, and CAM) as well as for specific species. For instance, the mortality rate of *A. fumigatus* in CAPA was determined providing insight into species-specific outcomes, which enabled a detailed understanding of mortality rates across the various SFIs and their respective causative pathogens. A survival analysis was conducted to explore the interplay between gender, age, and species-specific conditions. The Long-rank, Gehan, and Tarone-Ware tests assessed differences in survival patterns. The analysis also estimated median age at risk for each species-specific condition, by gender, using cumulative hazard functions and 95% CI. A binomial logistic regression model was employed to assess the risk of treatment outcomes with species-specific pathogens. The model calculated the probability of successful treatment outcome, accompanied by standard errors (SE) and Z-scores. These metrics enabled the evaluation of the likelihood of treatment success or failure in correlation with specific pathogens. To provide the optimal treatment strategy, a Crosstable for dependent outcome analysis was performed to predict mortality rates for the treatment outcome. This final step enabled the identification of the most effective treatment approach by examining the intersection of treatment options and mortality rates, providing actionable insights for clinicians to make data-driven decisions for the management of SFIs.

## Introduction

COVID-19 was recorded as the most severe pandemic calamity to humans of the current decade. Over 50 distinct COVID-19 strains have been documented worldwide. As of 14 July 2024, 775.7 million people had contacted the virus, leading to 7 million fatalities (“WHO Coronavirus (COVID-19) Dashboard” n.d.). The decline in death rate over the period was achieved by lockdown, social distancing measures, hygiene practices, quarantine of confirmed cases and vaccination (1,2). Alongside SARS-CoV-2 infection, secondary infections by fungi or bacteria increased the death rate. Yet, the COVID-19 treatment that includes antiviral, antibiotics, steroids, and immunosuppression drugs during prophylaxis medication controlled the bacterial infection, however not the fungal Infections (3). In addition to the compromised immune system, an individual with any of the following comorbidities: age, chronic obstructive pulmonary disease, diabetes mellitus, hypertension, steroids, immunosuppressive drugs (tocilizumab), severe lymphopenia, respiratory failure, malignancy, chronic liver disease and solid organ transplant were the supremacy factors for SFIs (4). As a preliminary means, the outbreak of invasive fungal infection in COVID-19 patients was driven by fungal spores’ inhalation (*Aspergillus* or *Mucorales*) or commensal to pathogen shift of *Candida* spp. Besides, COVID-19 patients exhibited reduced granulocyte activation, inactive phagocytosis, and down-regulation of antifungal cytokines from T-cell against *Aspergillus* spp. (5), *Mucorales* spp. and *Candida* spp.(6). Furthermore, the inadequate understanding of the fungal pathogenesis has been a major concern in prognosis during COVID-19. To our knowledge, the present review is the first of its kind to use meta-analysis and systemic review to examine the global surveillance of the SFIs associated with COVID-19 (from 2020 to 2024). The study emphasises on frequency of SFIs across the globe, species shift, gender-specific infection rates, the significance of medical history, the efficacy of steroid and antifungal, treatment outcomes (mortality rate), and fungal- polymicrobial mortality analysis.

### Global surveillance of SFIs in COVID-19 patients highlighting geographical patterns

A global meta-analysis on COVID-19 associated SFIs reported from 58 countries reveals, *Aspergillus* spp., *Candida* spp., and *Mucorales* spp. as primary species **(Figure 1.a)**. The systematic analysis ( Aug 2020 to May 2024) yielded 10,700 SFI cases with a male predominance (65.6 %) over females (34.3 %) and a mean age of 59.2 years (±14.5 SD). Of which, COVID-19 Associated Mucormycosis (CAM) was recorded 59.2 %, COVID-19 Associated Candidiasis (CAC: 21.6 %) and COVID-19 Associated Pulmonary Aspergillosis (CAPA: 19.1 %). From the reported countries, the notable population was observed from India (46.8 %), Italy (10 %), Iran (9.4 %), France (5.1 %), Spain (4.3 %), and Egypt (4.1 %) (**Figure 2.b)**. In specific, CAPA was diverse in 38 countries among the male (70.6 %) and female (29.3 %) population with a mortality rate of 58 % (95% CI). The leading nations are France (15.8 %), Spain (14.1 %), Italy (11.6 %), China (10%), Austria (7.25 %), United Kingdom (6.27 %), Turkey (3.8 %), Egypt (3.7 %), Netherlands (3.6 %) and Germany (3.5%). CAC was noted in 30 distinct nations causing a 54 % (95% CI) mortality rate among the population (Male: 59.6 %, Female: 40.3 %). Countrywide, Italy (36.1 %), Egypt (10.2 %), Iran (9.02 %), Spain (7.6 %), Greece (6.3 %), United States (5.17 %) and Turkey (4.7 %) comprised 80 % of all CAC cases. In CAM, India standalone accounted for 78 %, Iran (12 %), France (3.4 %) and Egypt (2 %) in a population of male (70.6%) and female (29.3%) with a mortality rate of 28 % (95% CI).

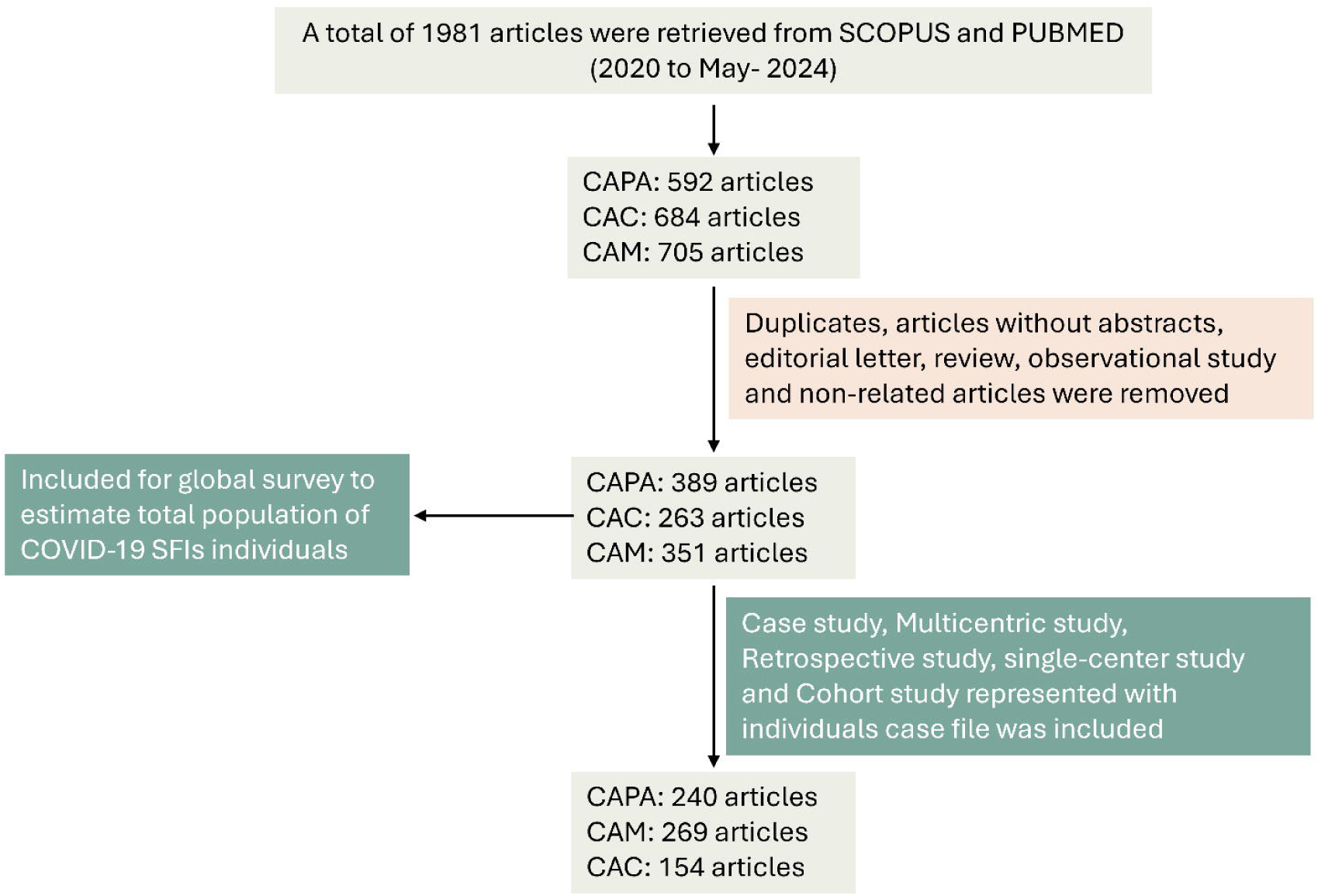

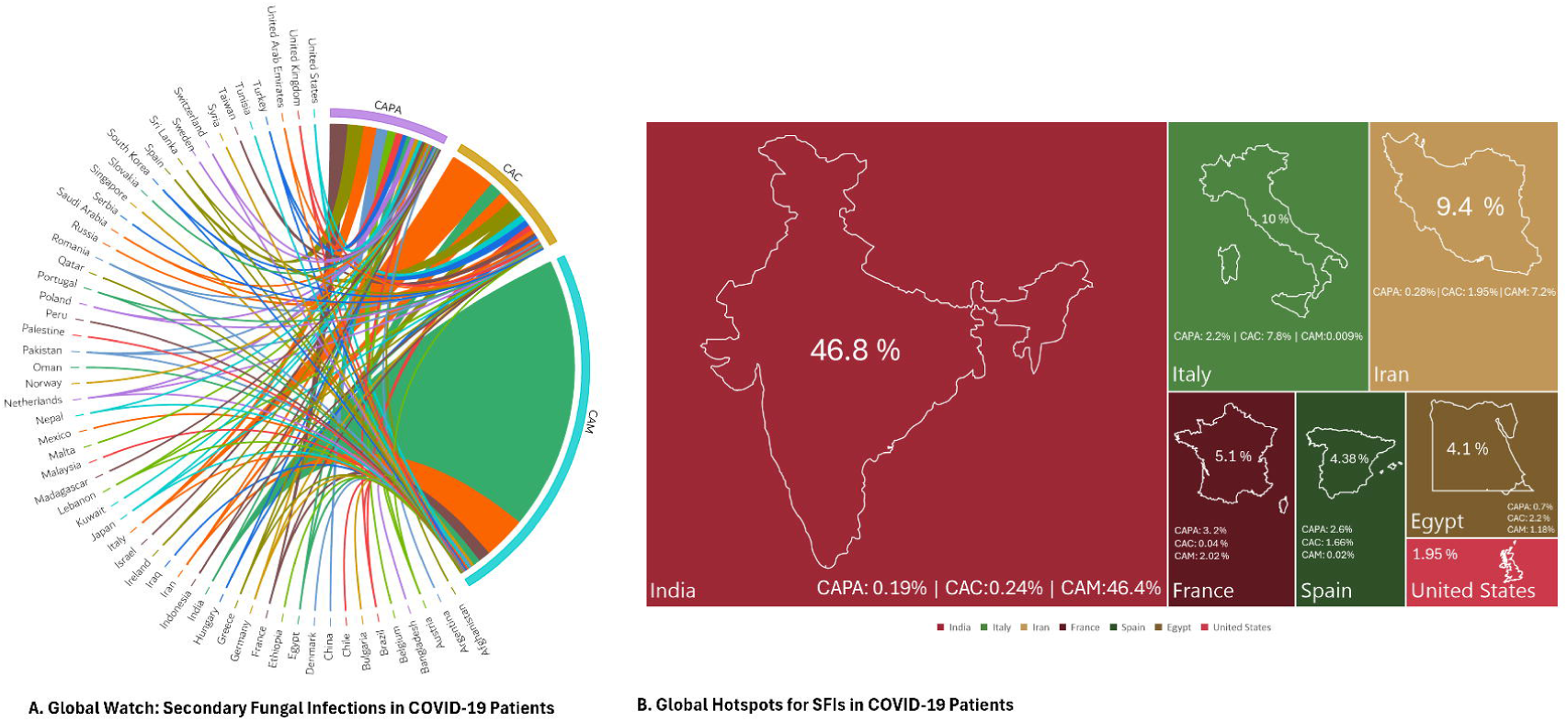

### Risk Factors, Species Shift, and Treatment Outcomes of SFIs

#### COVID-19 Associated Pulmonary Aspergillosis (CAPA)

Global surveillance on SFIs pathogens distribution reveals a 100% probability of *Aspergillus* spp. infections across 11 countries: Afghanistan, Argentina, Austria, Belgium, China, Israel, Kuwait, Malta, Norway, Slovakia, and Sweden with 95% Confidence Intervals (CI), indicating a high degree of certainty **(Figure 3)**. An emerging trend in eight countries - Saudi Arabia, Lebanon, South Korea, Indonesia, Ireland, Madagascar, Slovakia, and Sweden - has recorded a devastating 100% mortality rate (95% CI) **(Figure 4)**. Among the *Aspergillus* spp., *A. fumigatus* dominated with 57 % of the population causing a mortality rate of 55 % (95% CI). Concurrently, polymicrobial infections (26.4 %) and emergence of non-fumigatus spp. were noted consisting of *A. niger* (3.7 %), *A. flavus* (3.3 %), *A. penicillioides* (1.2 %), *A. nidulans* (0.8 %) *A. baumannii* (0.8 %), *A. quadrilineatus* (0.4 %), *A. ochraceus* (0.4 %), *A. calidoustus* (0.4 %), *A. terreus* (0.4 %) and *A. tubingensis* (0.4 %), confirmed at a 95% Cl **(Figure 5.a)**. The relative morality rates of non-fumigatus spp. of *A. niger* resulted in 33.3 % (9n), *A. flavus* (75 %;8n ), *A. nidulans* (50 %; 2n), *A. penicillioides* (100 %; 3n) and *A. baumannii* (50 %; 2n) with (95% CI). At the same time, 26.4% of CAPA patients had polymicrobial infections, drastically escalating the mortality rate to 100% (95% CI). The predominant polymicrobial infection was high in *A. fumigatus*, affecting 11.2% of the population, followed by *A. flavus* (6.4%), *A. niger* (4%), *A. tamarii* (0.8%), and *A. terreus* (0.4%) with 95% CI. The medical history in CAPA patients reveals a diverse range including diabetes mellitus (22.6 %), hypertension (15.4 %), respiratory diseases (14.3 %) heart-related diseases (10.6 %), without medical history (8 %), obesity (6.07 %), kidney related diseases (4.3 %), COPD (3.5 %), cancer (2 %), solid organ transplant (2 %), asthma (1.2 %) and IHD (1.1 %) **(Figure.7)** . Regardless of therapy, all CAPA patients with cancer and COPD had dismal mortality rates of 100 % and 85.7 % (Χ^2^_102_=121.64, P=0.092), respectively. The CAPA patients without pre-existing medical conditions had a higher mortality rate of 64.5% compared to diabetes mellitus (35 %), hypertension (33.3 %), Asthma (57.1 %). The comorbidities of diabetes mellitus with hypertension increased the mortality rate to 41.6%. In addition, CAPA patients with diabetes and multiple comorbidities, including OPD, obesity, hypertension, Asthma, cancer, hyperlipidemia, ischemic heart diseases, kidney related diseases, HIV, respiratory diseases, coronary artery disease, systemic sclerosis, polymyalgia rheumatica, vascular dementia, and ankylosing spondylitis HLA B27 conditions resulted in a 52.8% mortality rate . Hypertension paired with multiple comorbidities resulted in a 55.5% mortality rate, underscoring the critical need for managing complex health conditions. Antifungal treatment outcomes in CAPA patients revealed varying degrees of efficacy. Voriconazole was found to be a promising agent, achieving a 50 % survival rate. Notably, adjunctive dexamethasone therapy significantly enhanced survival to 68.4 %, demonstrating a synergistic effect. In contrast, combining Voriconazole with other steroids (corticosteroids, prednisolone and methylprednisolone) yielded a detrimental effect, resulting in a 75 % to 100 % mortality rate. Monotherapy with AmBisome was ineffective, with a 90 % mortality rate, and its combination with steroids did not improve outcomes. Conversely, the absence of both steroids and antifungal treatment was associated with a 78 % mortality rate. Thus, highlighting the complex interplay between treatment modalities and species specificity.

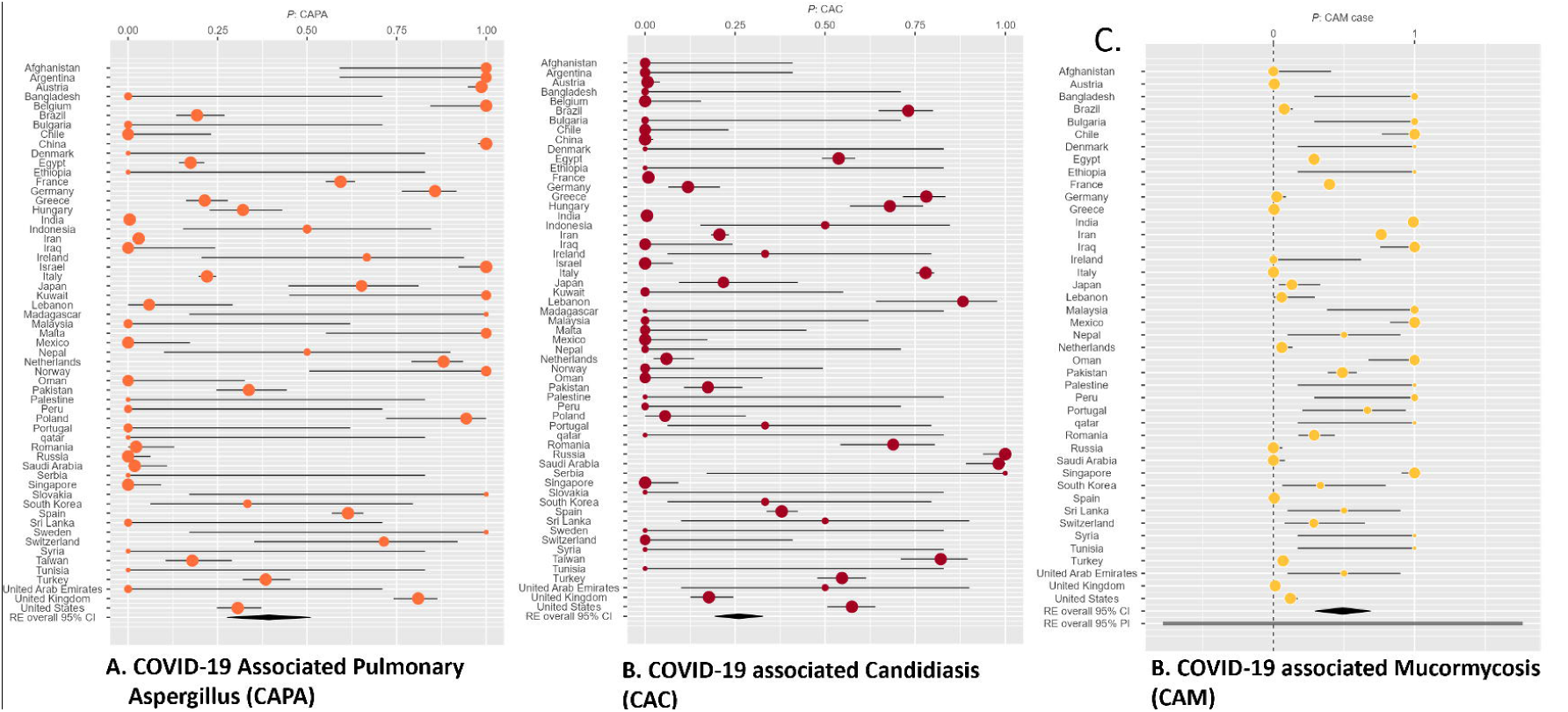

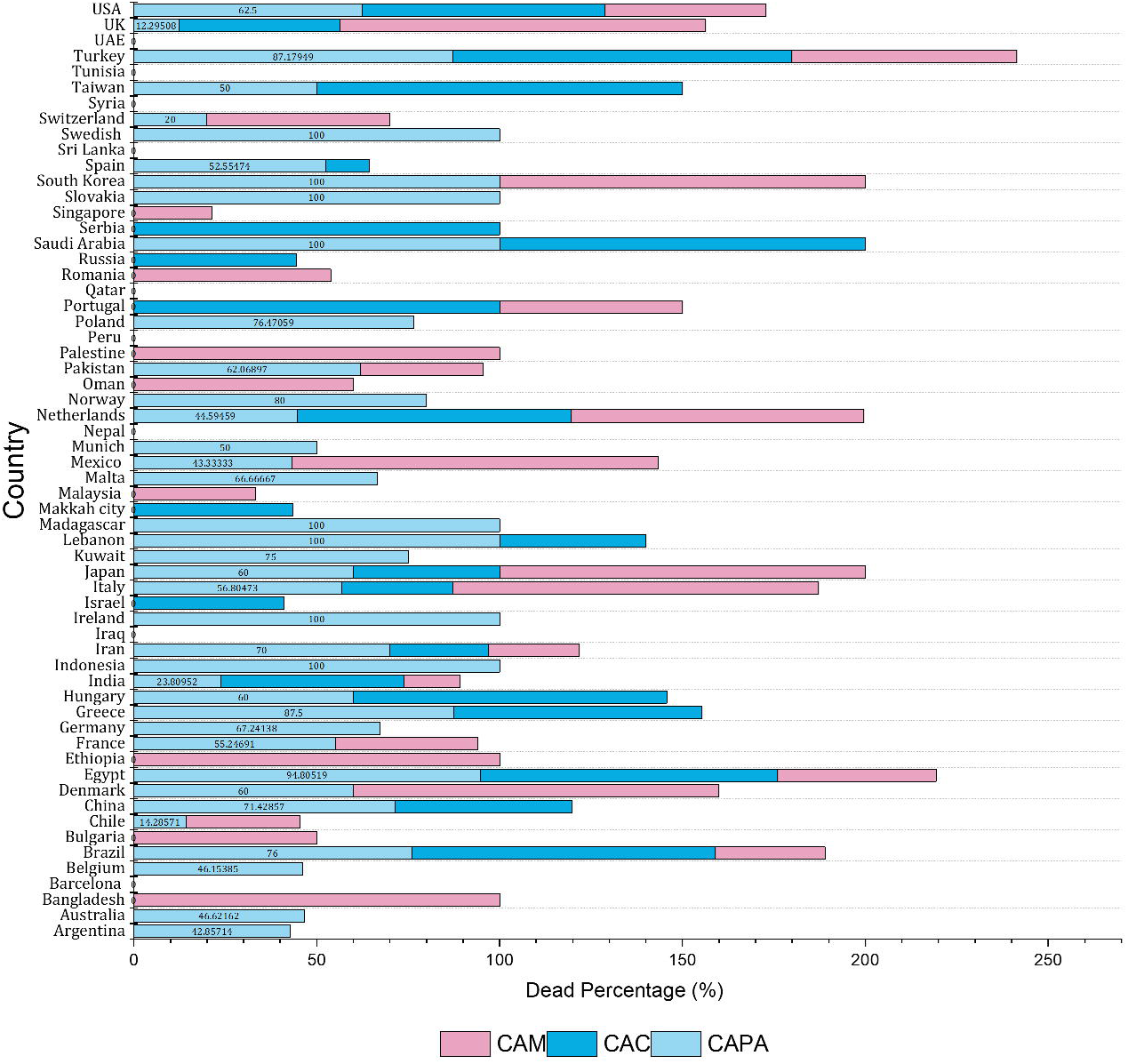

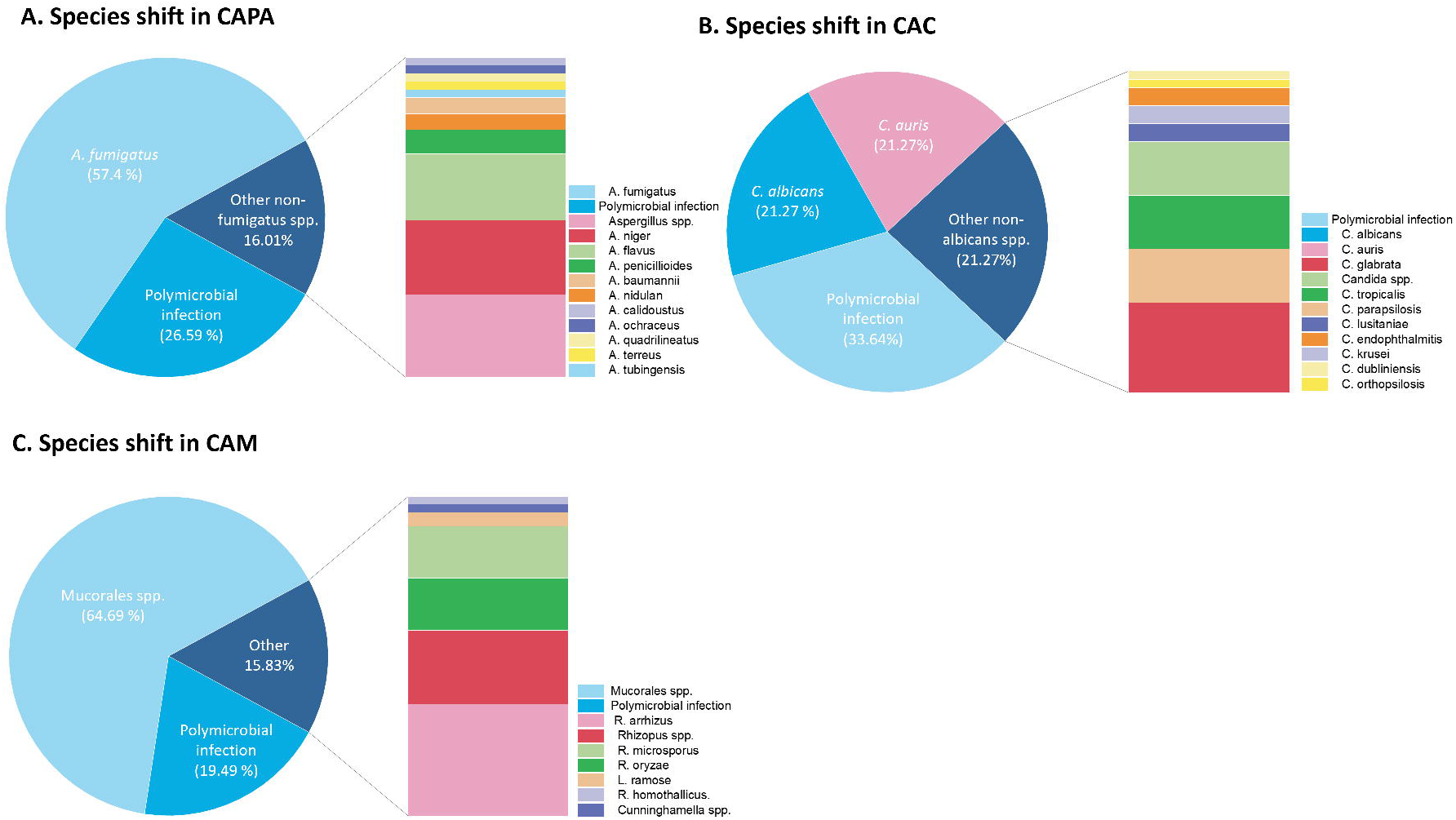

#### COVID-19-Associated Candidiasis (CAC)

A worldwide analysis of CAC in countries of Russia, Saudi Arabia, and Serbia reported a 100% incidence rate **(Figure 3.b)**. Subsequently, Saudi Arabia, Serbia, Taiwan, and Portugal exhibited a uniformly high mortality rate of 100%, whereas Russia demonstrated a divergent outcome **(Figure 4)**. In similar to CAPA, a prominent species shift of non-albicans spp. Over 40.26 % than *C. albicans* (20.77 %) was observed in CAC. The non-albicans spp. comprise of *C. auris* (20.7 %), *C. glabrata* (6.4 %), *C. parapsilosis* (3.8 %), *C. tropicalis* (3.8 %), *C. krusei* (1.2 %), *C. lusitaniae* (1.2 %), *C. endophthalmitis* (1.2 %), *C. orthopsilosis* (0.6 %), and *C. dubliniensis* (0.6 %) **(Figure 5.b)**. Simultaneously, the emergence of non-albicans spp. was associated with a statistically significant increase in mortality rates compared to *C. albicans*. The non-albicans spp. mortality rates rose, led by *C. krusei* and *C. dubliniensis* (100%; 2 each cases), followed by *C. glabrata* (60%; 10 cases), *C. tropicalis* (50 %; 6 cases, *C. lusitaniae* (50%; 2 cases ), *C. auris* (46.9%; 32 cases), *C. parapsilosis* (33.7%; 6 cases), and *C. endophthalmitis* (0%; 2 cases). The co-infection of *C. albicans* with non-albicans spp. of *C. tropicalis, C. glabrata, C. parapsilosis* increased the death probability up to 100 % (95% CI). Among these, polymicrobial infections with *C. auris* accounted for 18.8% followed by *C. albicans* (9.7 %), *C. glabrata* (1.9 %) and *C. parapsilosis* (1.9 %). The polymicrobial infections involving non-albicans spp. exhibited higher mortality rates compared to *C. albicans* with 95% CI. Specifically, *C. glabrata*, *C. auris*, and *C. parapsilosis* demonstrated significantly higher mortality rates - 100% (3 cases), 75.8% (29 cases), and 66.6% (3 cases), respectively. In contrast to *C. albicans*, which had a mortality rate of 33.3% (15 cases). Interestingly, coinfection of *C. auris* with *C. albicans* was not identified. The co- infection *C. auris* with *C. glabrata* and other bacterial spp. are observed **(Figure 6.b)**. In case of *C. albicans*, the polymicrobial infection was very diverse among bacteria and fungi spp. The risk factors associated with CAC include hypertension (25.4 %), diabetes mellitus (24.1%), catheterization (12.0 %), ischaemic heart disease (9.32 %), cancer (7.2 %), chest related disease (5.2 %), COPD (5.2 %), obesity (3.9 %), hypothyroidism (3.7 %), kidney failure (3.7 %), and asthma (1.05 %) **(Figure 7).** A significant correlation between comorbidities and increased mortality rates was observed in CAC patients, mirroring findings in CAPA. Specifically, multiple comorbidities accompanying diabetes mellitus increased mortality from 50% (n=8) to 62.7% (n=51). A similar pattern emerged for individuals with multiple comorbidities associated with catheterization, where mortality rates rose from 20% (5 cases) to 60% (15 cases), and for those with hypertension, from 27.2% (11 cases) to 64% (24 cases). The first-line practice of caspofungin in combination with corticosteroids increased the survival rate from 28.5 % to 57 %, followed by corticosteroids and micafungin combination (50 %).

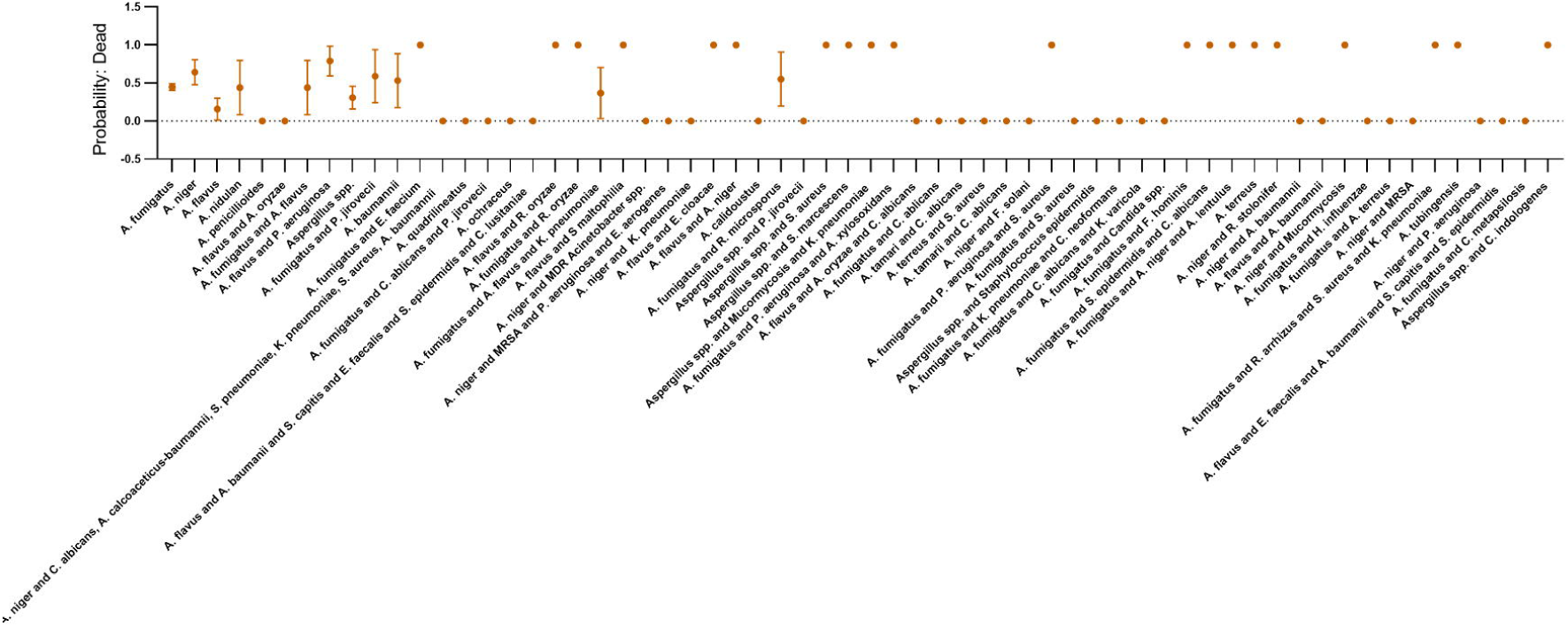

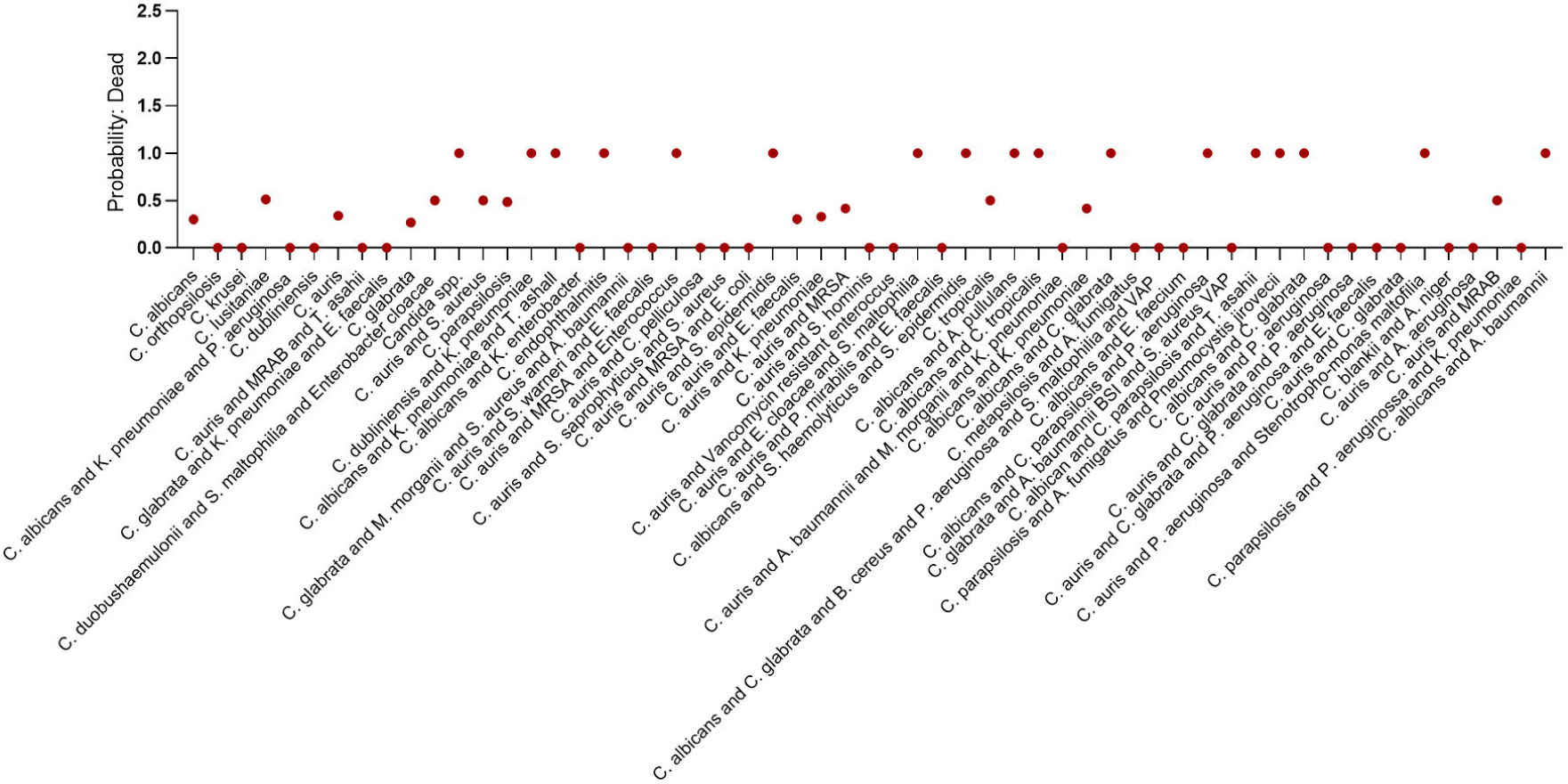

#### COVID-19-Associated Mucormycosis (CAM)

The global trends of CAM infection were exclusively reported (100%) in 13 countries: Bangladesh, Bulgaria, Chile, Denmark, India, Malaysia, Mexico, Palestine, Peru, Qatar, Singapore, Syria, and Tunisia **(Figure 3.c)**. Of which, the United Kingdom, Japan, Italy, South Korea, Denmark, Mexico, and Bangladesh countries resulted in a 100 % mortality rate **(Figure 4)**. Diabetes mellitus emerged as the defining trademark of CAM patients, with a prevalence of 79.8 %. Subsequent with hypertension (10.4 %), and other (3.7 %) includes kidney transplantation, dyslipidemia, coronary artery disease, chronic kidney disease, ischemic heart disease. A comparative analysis of mortality rates revealed significant differences based on underlying medical conditions. Specifically, individuals with diabetes mellitus alone exhibited a mortality rate of 27% (140 case), whereas those without pre-existing conditions had a mortality rate of 32.2% (59 cases). Hypertension was associated with a substantially higher mortality rate of 60% (5 cases). In contrast, individuals with comorbid diabetes mellitus and hypertension demonstrated a relatively lower mortality rate of 18.5% (27 cases). The global CAM burden was predominantly driven by *Mucorales* spp. (66.9%). Other notable causative agents included *Rhizopus arrhizus* (15%), *Rhizopus* spp. (10%), *Rhizopus oryzae* (7%), and *Rhizopus microsporus* (7%) **(Figure 5.c)**. Species-specific mortality rates demonstrated significant variability of *Mucorales* spp. resulting 19.3 % (176 cases) followed by *R. microsporus* (28.6 %; 7 cases), *R. arrhizus* (33.3%; 15 cases), *R. oryzae* (42.9 %; 7 cases), *L. ramose* (50%; 2 cases) and *Rhizopus* spp. (60 %; 10 cases). A notable outbreak of treatment failure occurred in CAM with polymicrobial infections, resulting in a marked escalation of mortality rates. For instance, *Rhizopus* spp. associated mortality rate surged from 60 % to 100 % upon coinfection with *Aspergillus* spp., *P. aeruginosa*, *S. maltophila*, *C. glabrata*, *B. bronchisepticum* and *P. jirovecii*. A comparable scenario calls for polymicrobial infections of *R. microsporus, R. arrhizus, L. ramose,* and *R. microsporus* **(Figure 6.c).** In response to treatment, the practice of AmBisome followed by azole drug was effective in all pre-medical conditions (diabetes mellitus, hypertension, etc.) **(Figure 6.c).** The combination of antifungal agents with steroids did not exhibit synergist and antagonist effects.

### Insight into gender and age related to SFIs

In terms of gender and age, the survivability rates decline with advancing age in both genders. Notably, median age varies by gender and fungal species. In *Mucorales* spp. the median age of Male (60) and Female (68); *C. albicans*; Male (69) and Female (67); non-albicans spp.; Male (72), Female (82); *A. fumigatus*; Male (73) and Female (75); non-fumigatus spp.; Male (70) and Female (79). Interestingly, the polymicrobial infections increased the median ages across various fungal species, except for non-albicans spp. Specifically, in *Mucorales* spp., Male (71) and Female (73), *A. fumigatus*; Male (74) and Female (68), non-fumigatus spp.; Male (73) and Female (69), *C. albicans*; Male (77) and Female (75) and non-albicans spp.; Male (64) and Female (74) **(Figure 8)**.

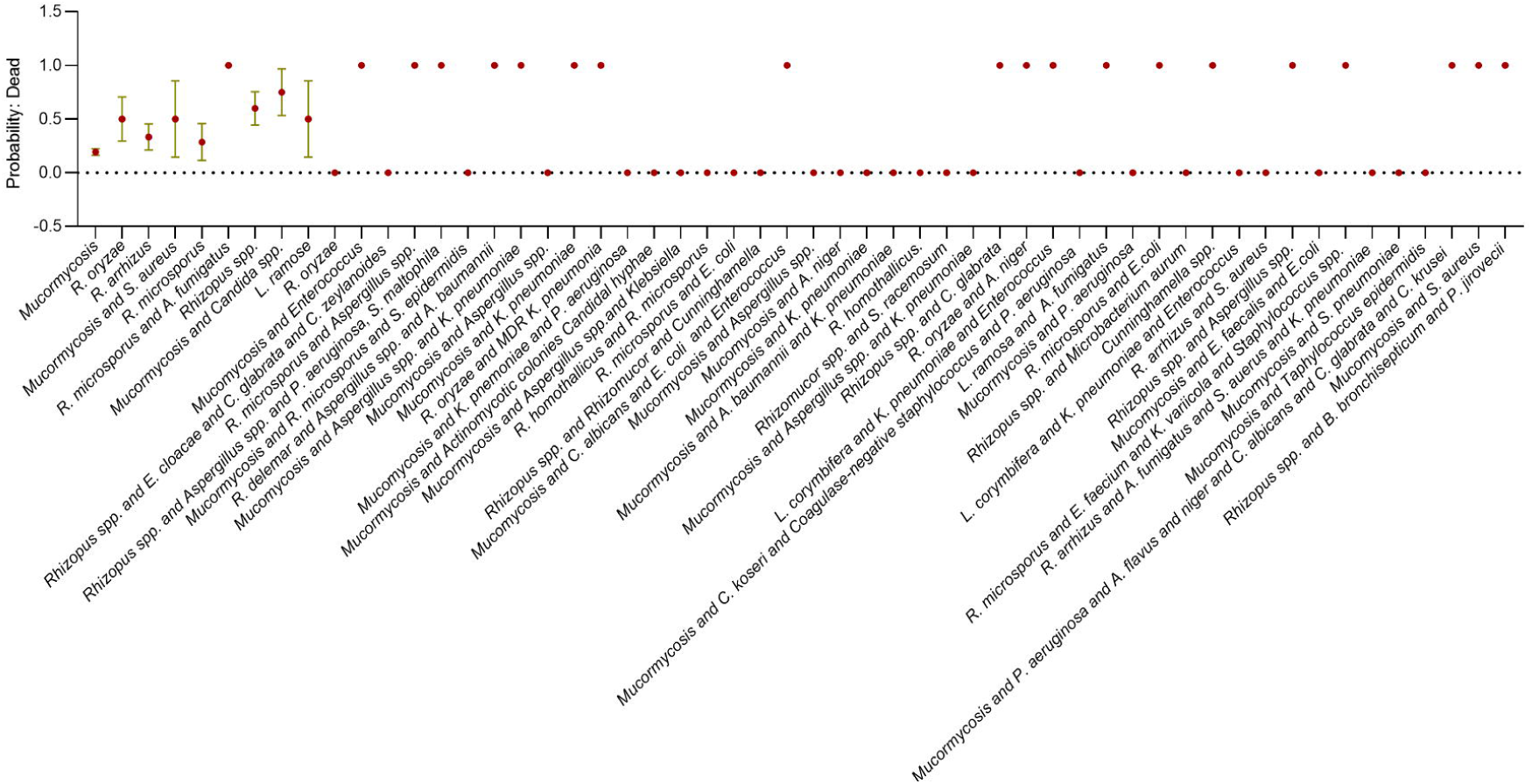

### Fungal virulence Attribution and the Severity of COVID-19

The global meta-analyses on SFIs associated with COVID-19 revealed a predominance of specific fungal pathogens, notably *Mucorales* spp., *Aspergillus* spp., and *Candida* spp. Whereas species-specific virulence factors led to the dominance of *A. fumigatus* in CAPA, *C. auris* in CAC, and *Mucorale*s spp. in CAM. A common virulence trait among these dominant pathogens was thermotolerance. *A. fumigatus* exhibits remarkable thermotolerance, remaining viable up to 41°C. In contrast, *A. flavus* shows decreased germination by 45% at this temperature, while *A. niger* exhibits complete inhibition of germination (7). The constitutive overexpression of heat shock protein HSP90 in *C. auris* favours growth up to 42 °C differing from another *Candida* spp. (8). Similarly, the dormant spores of *Rhizopus* spp. withstand temperature up to 50 °C (9). Thus, in COVID-19 patients with fever (>38 °C) do not hinder *A. fumigatus*, *C. auris* and *Rhizopus* spp. developing SFIs (10). Furthermore, the alkaline protease produced by *A. fumigatus* during COVID-19 infection mediates the proteolytic cleavage at S1/S2 junction of the viral spike protein. Thereby promoting the endocytic uptake of pseudo virions (11). In the case of CAM, *Mucor lusitanicus* genome has identified 17 CotH-like proteins. Where CotH8 and CotH 17 enhance the macrophage interaction. CotH7 binds with integrin α3β of alveolar epithelial cells mediating the *Mucorales* spp. invasion (12). Similarly, CotH2 and CotH3 bind with Glucose-Regulated Protein 78 (GRP78) expressed on endothelial cells regulating the invasion of *Rhizopus* spp. (12). Also, GRP78 acts as a co-receptor for SARS-CoV-2 spike protein regulating virus entry playing a crucial role in CAM (13,14). In addition, the diabetes mellitus condition increases the GRP78 expression leading to CAM susceptibility (15). Following, *Mucorales* spp. genome possesses an elaborate iron scavenging machinery, leveraging iron from haemoglobin, deferoxamine and circulating free irons. The intricate system is mediated by 13-siderophore permeases, 6-copper oxidases, 3-ferric reductases, 2-heme oxygenase, 1-high affinity permease (FTR1), 1-SreA and 1-ferritin. Of which, the FTR1 gene in *Rhizopus* spp. is the crucial virulence factor regulating the transport of iron across the cell membrane (16). And highly expressed in diabetic ketoacidosis (DKA) condition (16). During COVID-19 infection, the coronavirus causes hemoglobin disruption releasing the serum iron level up to 842.09 ng/mL (17). In parallel, ferritin is reimbursed to reduce free iron in serum (18). During this course of time, the individual was exposed to *Mucorales* spp. develops CAM (19). As such, compared to *A. fumigatus* and *C. albicans*, the *Rhizopus* spp. incorporate the iron 8-fold and 40-fold higher respectively (20). In case of CAC, COVID-19 patients receiving immunosuppression drugs and broad-spectrum antibacterial disrupt the gut microbiota, reduce the proliferation of immune cells (th17 cells,CD4 T-cells), mucosal tissue damage and expression of phospholipase/protease enzyme in *Candida* spp. leading to systemic infection (21–25). Furthermore, Candida’s adhesion properties facilitate dense biofilm formation on catheter surfaces, significantly increasing the risk of catheter-related bloodstream infections (26,27). Our meta-analysis substantiates this link, emphasizing the clinical significance of Candida biofilms in C-RBSI.

## Discussion and Conclusion

To our knowledge, this is the first kind of study that involves a systematic review and a global meta-analysis on SFIs associated with COVID-19. SFIs in COVID-19 have alerted the emergence of fungal pathogens. In association with COVID-19, the existence of *Mucorales* spp., *Aspergillus* spp., and *Candida* spp., was reported from 39, 38, and 31 countries respectively. Among which, CAM accounted for a high population with less mortality rate of 28 %. Whereas the significantly smaller population of CAPA and CAC resulted in high mortality rates of 58% and 54 % respectively. In individuals with diabetes, particularly those experiencing ketoacidosis, an inherited ketone reductase system enables *Mucorales* spp. to flourish in glucose-rich environments. Furthermore, diabetic ketoacidosis enhances the expression of iron scavenging systems, allowing them to exploit iron from haemoglobin, myoglobin, and coronavirus-induced haemolysis. This synergistic relationship between diabetes and iron scavenging promotes the growth and pathogenesis of *Mucorales* spp. in of CAM. The enhanced adhesion and biofilm properties of aided *Candida* spp. in conquering the catheterized individuals. The propensity of *Candida* spp. to form robust biofilms and adhere to surfaces has made them a significant threat to catheterized individuals and female susceptible to vulvovaginal infections (28). Furthermore, individuals with cancer, ischemic heart diseases, obesity and hypothyroidism also exacerbate the risk of developing CAC through unknown mechanisms. In cases of CAPA, the strong adherence properties of conidia of *Aspergillus* spp. along with efficient switching to filament form in the presence of limited energy resources has driven the *Aspergillus* spp. infection associated with individuals with respiratory diseases (29). Subsequently, the meta-analysis reveals individuals without pre-medical conditions are also highly susceptible to CAPA followed by CAC and CAM. In terms of age distribution, males showed a lower median age (60 years) susceptible to the SFIs compared to females (67 years). Polymicrobial infection has been observed in older age groups with >65 years. During treatment in CAC, both caspofungin and micafungin in combination with corticosteroids worked well. Similarly, the combination of Voriconazole with Dexamethasone was effective in CAPA. In CAM, AmBisome, Fluconazole, and Itraconazole functioned independently without steroids.

Furthermore, global fungal genomic surveillance is required to understand geographic diversity, the evolution of new strains, phylogenetic analysis, genetic variations, pan-genome analysis, virulome and resistome. Deciphering these analyses provides insights into this pathogen evolution which is crucial for understanding disease management and suggests potential targets for therapeutic interventions.

## Data Availability

All data produced in the present work are contained in the manuscript

## Acknowledgments

The authors wish to express their deepest thanks to the research community for their tireless efforts in investigating secondary fungal infections linked to COVID-19, thereby facilitating the completion of this manuscript. The authors express their gratitude to SASTRA Deemed University for providing infrastructural facilities and financial support.

## Author Contributions

AM performed all the experiments and analysed the results. SR performed data extraction for CAM. Manuscript was written and verified by AM and JR.

---

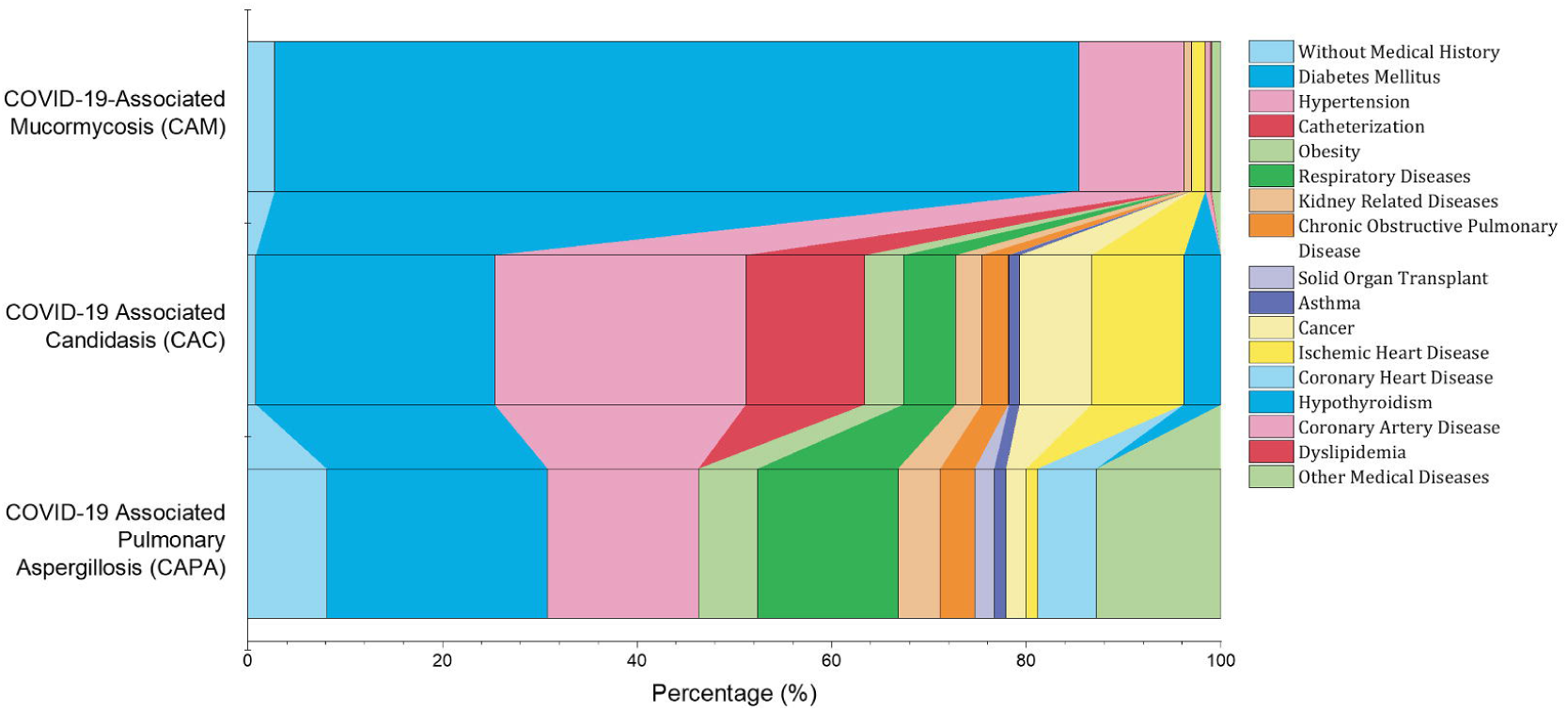

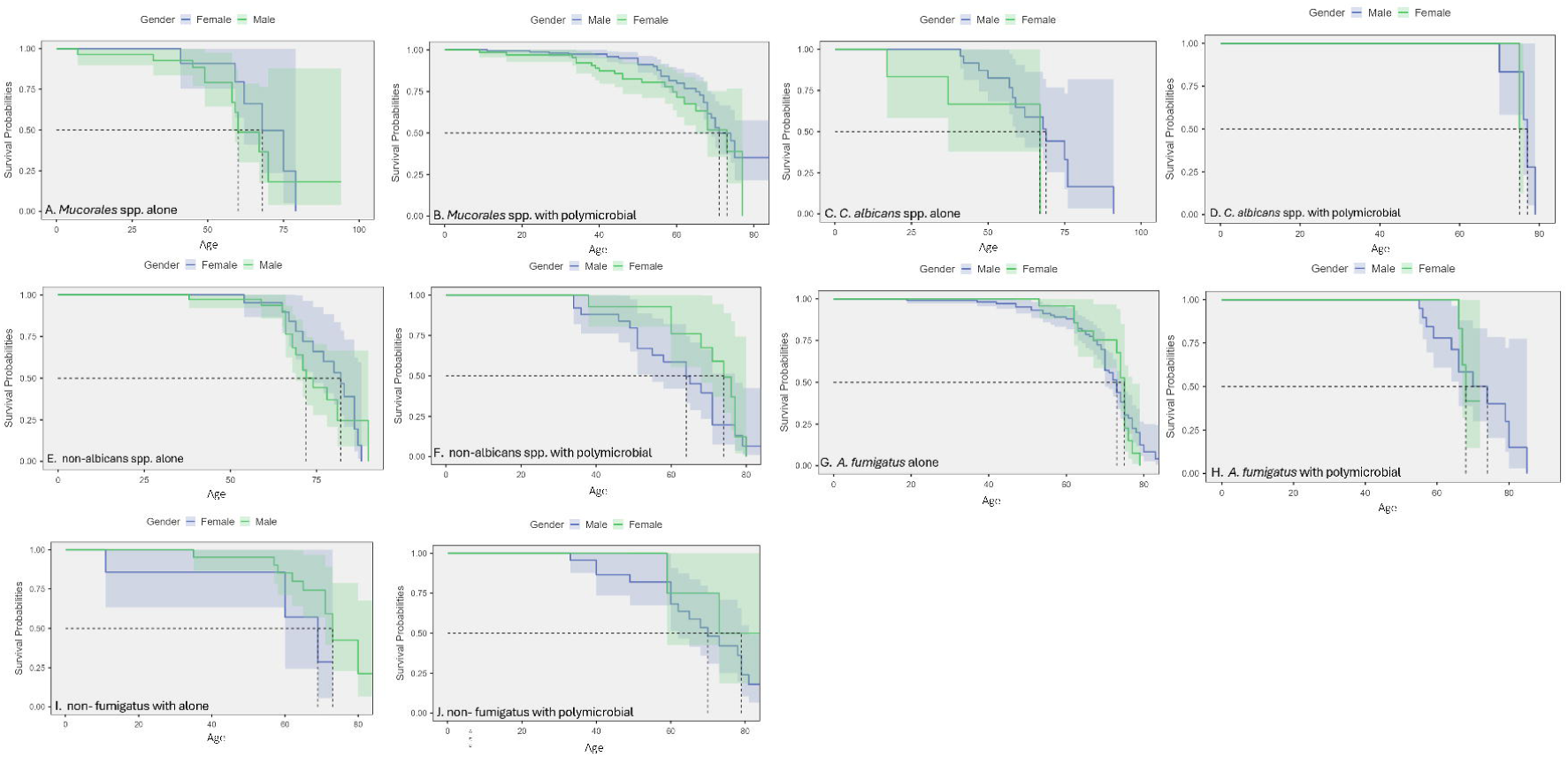

